# The predictive capacity of polygenic risk scores for disease risk is only moderately influenced by imputation panels tailored to the target population

**DOI:** 10.1101/2023.08.29.23294769

**Authors:** Hagai Levi, Ran Elkon, Ron Shamir

## Abstract

Polygenic risk scores (PRS) predict individuals’ genetic risk of developing complex diseases. They summarize the effect of many genetic variants discovered in genome-wide association studies (GWASs). However, to date, large GWASs exist primarily for the European population and the quality of PRS prediction declines when applied to target sets of other ethnicities. A key step in using a PRS is imputation, which is the inference of un-typed SNPs using a set of fully-sequenced individuals, called the imputation panel. The SNP genotypes called by the imputation process depend on the ethnic composition of the imputation panel. Several studies have shown that imputing genotypes using a panel that contains individuals of the same ethnicity as the genotyped individuals improves imputation accuracy. However, until now, there has been no systematic investigation into the influence of the ethnic composition of imputation panels on the accuracy of PRS predictions when applied to ethnic groups that differ from the population used in the GWAS.

In this study we estimated the effect of imputation of the target set on prediction accuracy of PRS when the discovery (GWAS) and the target sets come from different ethnic groups. We analyzed twelve binary phenotypes and three populations from the UK Biobank (Europeans, South-Asians, and Africans). We generated imputation panels from several ethnic groups, imputed the target set using each panel, and generated PRS to compute individuals’ risk scores. Then, we assessed the prediction accuracy obtained from each imputation panel. Our analysis indicates that using an imputation panel matched to the ethnic group of the target population yields only a marginal improvement and only under specific conditions. Hence, while a target-matched imputation panel can potentially improve prediction accuracy of European PRSs in non-EUR populations, the improvement is limited.

## Introduction

Polygenic risk scores (PRSs) play a prominent role in the vision of precision medicine, offering significant potential for enhancing healthcare outcomes. They can predict individuals’ risk of developing a certain disease based on summary statistics of genome-wide association studies (GWASs). As ethnic groups differ in genetic structure, the set of individuals included in a GWAS, also called the **discovery set**, are usually from the same ethnicity. The PRS constructed from GWAS can then be used to calculate risk scores for another group of individuals, also called the **target set**. Ideally, the discovery and the target sets should come from the same ethnic group. The prediction accuracy of PRS decreases as the genetic distance between the discovery and the target set increases (Martin *et al*., 2019). At present, most GWASs have been compiled from the European (EUR) population (Martin *et al*., 2019), and applying the PRSs generated from them to non-EUR individuals has lower accuracy. Therefore, the health benefits that can be achieved using current PRS models are still not effectively applicable to most of the world’s population.

Genetic profiling of individuals in the discovery and target sets is usually obtained by SNP arrays that genotype a predefined subset of several hundreds of thousands of SNPs per individual. Nevertheless, additional information is often contained in millions of other SNPs that were not genotyped. Furthermore, discovery and target sets are often genotyped by different arrays. Computationally calling non-genotyped SNPs is done using **imputation**, a family of methods that infer untyped SNPs using a set of fully-sequenced individuals for whom several millions of SNPs are called (Yun *et al*., 2009; Chen *et al*., 2020). That set of individuals is called the **imputation panel**. Imputation methods rely on the fact that chromosome recombination during evolution has generated genomic intervals within the human genome where SNP alleles tend to be inherited together (a phenomenon called Linkage disequilibrium (LD) patterns). Imputation methods use the panel to complete untyped SNPs by assuming similar SNP correlation patterns (haplotype patterns) for the individuals in the target (genotyped) and the panel (sequenced) sets (Howie *et al*., 2009). Most currently available imputation panels consist of predominantly EUR individuals (e.g., HRC (McCarthy *et al*., 2016)) or EUR mixed with other populations (e.g., 1000 Genomes (Auton *et al*., 2015), HapMap (Rotimi *et al*., 2007) and TOPMed (Taliun *et al*., 2021)). Therefore, properties of the genetic structure (e.g., LD structure and minor allele frequency (MAF)) might differ between these panels and non-EUR populations. Studies have shown that using an ethnic-matched imputation panel improves the accuracy of completing untyped SNPs in the genotyped individuals (Carmi *et al*., 2014; Ahmad *et al*., 2017; Lencz *et al*., 2018). Nevertheless, accurately completing untyped SNPs does not necessarily imply better risk prediction with PRS. As of now, there has been no systematic testing of the effectiveness of matching the ethnicities of the imputation panel and the target set to enhance PRS risk prediction.

In this study, we estimated the effect of the ethnicities of the imputation panel and the target set on the prediction accuracy of PRS in cases where the discovery and the target sets come from different ethnic groups. We analyzed 12 binary phenotypes and three populations from the UK Biobank (UKB) (Europeans, South-Asian, and Africans). We generated imputation panels from several ethnic groups, imputed the target set according to each panel, used PRS to compute individuals’ risk scores, and finally, compared the performance of risk prediction among the different imputation panels.

## Results

We started by evaluating the quality of imputation as a function of the ethnic composition of the imputation panel and the imputed individuals. For a given imputation panel and a test set of fully sequenced individuals, we masked all SNPs in the test set except those included in the UKB genotyping chip (n=784,849) (Bahcall, 2018). Then, we imputed the masked SNPs using the imputation panel, and computed the fraction of the SNPs that were correctly imputed. We call that fraction the **imputation accuracy**.

To generate imputation panels for different ethnicities, we used the 1000 Genomes Project (1KG) dataset (Auton *et al*., 2015), which contains sequences of ∼2500 individuals annotated by their country (subpopulation) and continent (super-population) of origin. For each super-population, we took five groups of 70 individuals from each of its five subpopulations to generate an imputation panel (n=350). Using these sets, we generated an imputation panel for each super-population: EUR, East Asians (EAS), Africans (AFR) and South Asians (SAS). Similarly, for each super-population, we generated a test set by collecting 100 individuals from it, 20 from each subpopulation (Table S1). In addition, we generated another target set of two African subpopulations from the 1kG that do not appear in the African imputation panel (n=145; AFR2; Table S1). The target sets and the imputation panels were disjoint. After masking SNPs that do not appear on the UKB chip, we imputed each target set using the different imputation panels we generated and calculated the imputation accuracy. This test was conducted for each autosomal chromosome separately.

Table 1 shows the average imputation accuracy on different populations when using imputation panels of different ethnicities. The imputation accuracy deteriorates as the genetic distance (*F*_*st*_ ; calculated using (Bhatia *et al*., 2013); Table 2) between the ethnic groups increases. The genetic distance can also be visualized using a PCA of the SNPs (Figure 1). For example, the closest super-population to EUR is SAS (*F*_*st*_ *=* 0.03), then EAS (0.1), and the farthest is AFR (0.12). Accordingly, imputation accuracy obtained on the EUR test set was 0.99, 0.982, 0.957 and 0.947 using EUR, SAS, EAS, and AFR imputation panels, respectively. The agreement between distance and imputation accuracy also holds for other super-populations and on each chromosome individually (Table S2).

**Table 1.**
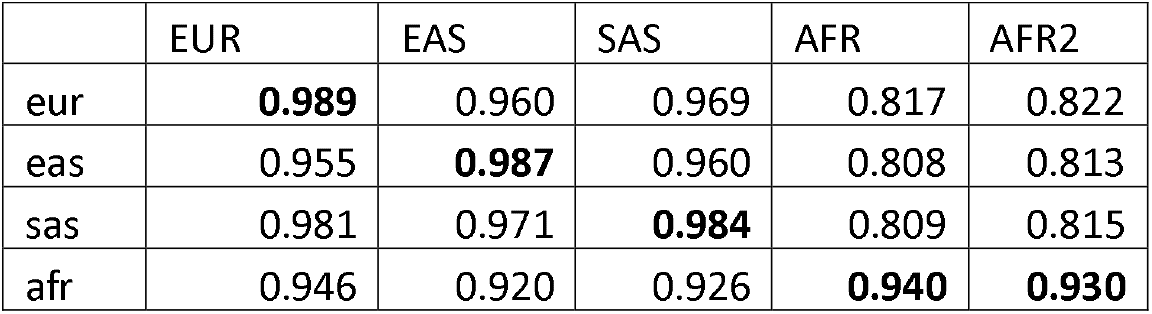
Accuracy of imputing different populations using panels from different ethnicities. Columns: The imputed population. Rows: The ethnicity of the imputation panel. The numbers are the accuracy averaged across chromosomes. (Best performing panel for each test set is in bold)

|  | EUR | EAS | SAS | AFR | AFR2 |
| --- | --- | --- | --- | --- | --- |
| eur | **0.989** | 0.960 | 0.969 | 0.817 | 0.822 |
| eas | 0.955 | **0.987** | 0.960 | 0.808 | 0.813 |
| sas | 0.981 | 0.971 | **0.984** | 0.809 | 0.815 |
| afr | 0.946 | 0.920 | 0.926 | **0.940** | **0.930** |

**Table 2.**
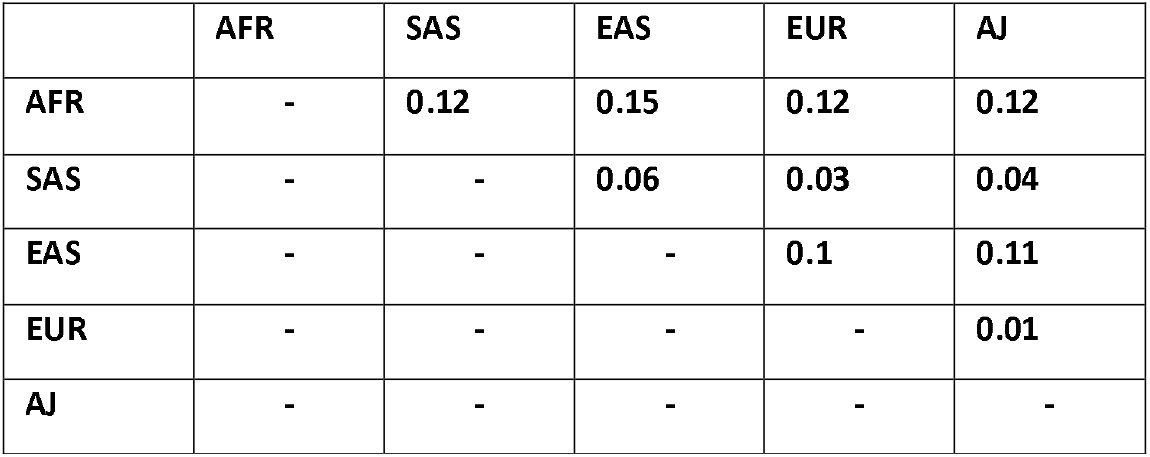
Genetic distance (Hudson’s *F*_*ST*_) between ethnic groups. AJ: Ashkenazi Jewish.

**Figure 1.**
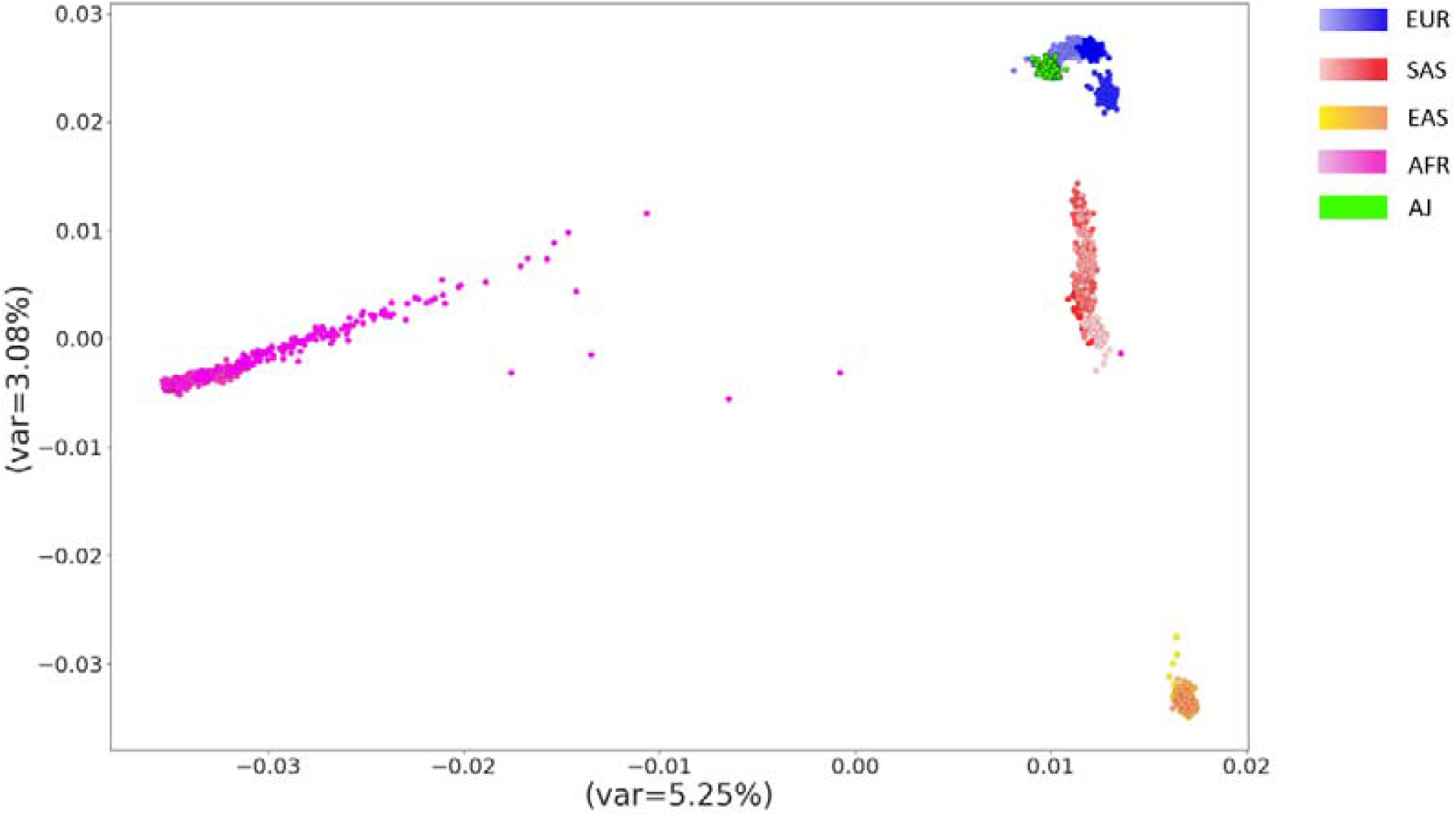
Principal Component Analysis on genotypes of five super-populations from the 1000Genomes project (n=2157, Bergström et al., 2020) and Ashkenazi Jews (n=128 (Carmi et al., 2014)), demonstrating genetic distances between populations (EUR: European; EAS: East Asians; SAS: South Asians; AFR: Africans; AJ: Ashkenazi Jews)

Two additional observations emerged from this analysis: (1) The highest accuracy on African individuals - obtained using the AFR imputation panel - was substantially lower than other ethnic groups. For example, the accuracy obtained on the AFR and AFR2 sets was 0.94 and 0.93, respectively, whereas, for the EUR, EAS, and SAS sets, when using ethnically-matched imputation panels, the accuracy was 0.989, 0.987 and 0.984, respectively. (2) An AFR imputation panel can be used for imputing EUR genotypes with a limited decrease in accuracy (0.989 vs. 0.946). However, the decrease in the other direction is much larger: using a non-AFR panel for imputing AFR individuals causes a large drop in the imputation accuracy (e.g., 0.94 using the AFR panel vs. 0.817 using the EUR panel). These findings reflect the more complex genetic structure of the AFR population compared to non-AFR populations.

Next, to include in our analyses a population that is more closely related to EUR, we added a cohort of fully sequences Ashkenazi Jewish (AJ) individuals from (Carmi *et al*., 2014), and conducted a similar analysis on this ethnic group. We generated an AJ imputation panel from 100 individuals and tested the imputation accuracy on a disjoint set of 27 individuals from the same study. In addition, we generated three imputation panels of 100 individuals each from each of the EUR, EAS AFR populations from the 1KG and used them to impute the same 27 AJ genotypes. Generally, the results of this analysis (Tables 3, S3) were consistent with the results presented above. The only exception was that the AFR imputation panel did slightly better than the EAS imputation panel. Notably, both populations are very far from the AJ (*F*_*st*_ = 0.11 and 0.12 for EAS and AFR, respectively).

**Table 3.** Imputation accuracy on an Ashkenazi Jewish (AJ) test set using panels from different ethnicities, and the genetic distance (Hudson’s *F*_*ST*_) between AJ to the other ethnic groups.

|  | Imputation accuracy | Genetic distance |
| --- | --- | --- |
| AJ | 0.956 | - |
| EUR | 0.951 | **0.01** |
| EAS | 0.917 | **0.11** |
| AFR | 0.928 | **0.12** |

The analyses above suggest that imputing genotypes using an ethnic-matched panel is more accurate. Next, we sought to examine the effect of the imputation panel on the performance of PRS prediction when applied to target sets of different ethnicities than the discovery (GWAS) population. We used UKB data and focused on three super-populations: EUR (n= 472,694), SAS (n=9,881) and AFR (n=8,060), and 12 diseases that had sufficient representation of SAS and AFR cases (Table S4; Methods). For each disease, we ran GWAS on the EUR population from the UKB, created a PRS and tested its prediction quality on the SAS and AFR target sets imputed using the three imputation panels that we generated previously from the three super-populations in the 1KG project: EUR, SAS, and AFR.

We applied three methods for PRS construction - P+T with discovery-set LD, P+T with target-set LD and Lassosum - and evaluated PRS performance using a nested cross-validation scheme (see Methods). We calculated the odds ratio per 1 unit of standard deviation (OR per 1SD) to measure the quality of the PRS prediction. We also evaluated the performance of PRS that were built only on SNPs directly typed by the SNP array (i.e., without imputed SNPs). Figure 2 summarizes the results on Lassosum, the best performing PRS method.

**Figure 2.**
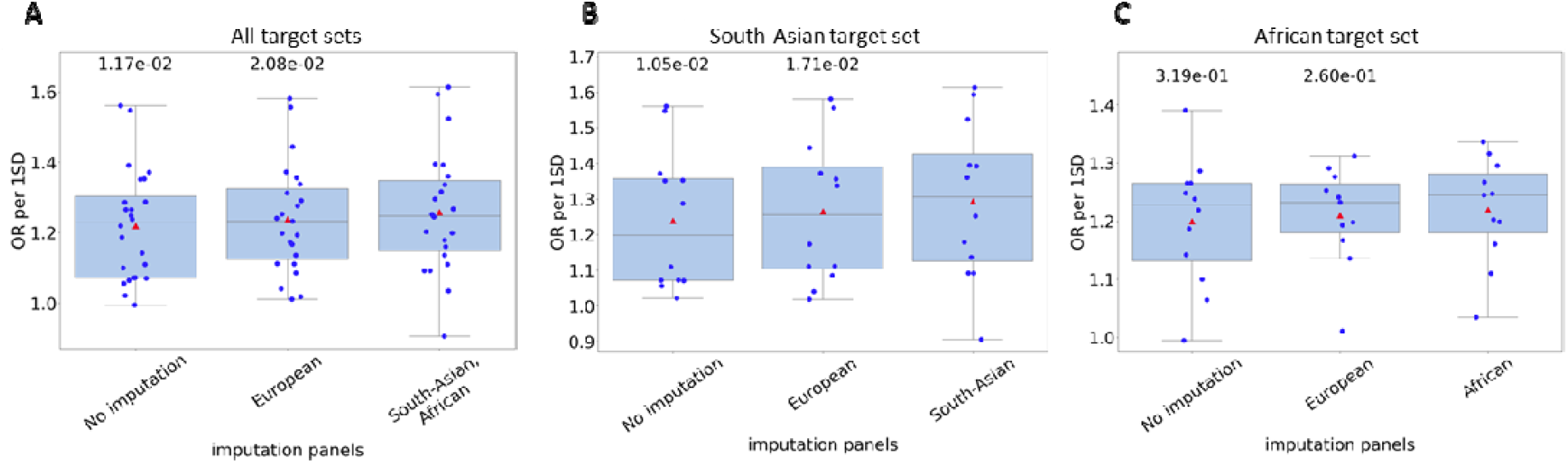
The effect of ethnic composition of the imputation panel on the performance of PRS constructed by the Lassosum method when applied to target set of different ethnicity than the GWAS. The graphs show OR per 1SD of 12 traits. PRSs were built from GWASs computed on UKB EUR individuals. Results are shown for (A) both SAS and AFR (B) SAS only (C) AFR only. The p-values above each boxplot compare the results with the imputation panel listed below it to the results with the imputation panel of the target population. P-values were calculated using one-tailed Wilcoxon test. Red triangles are the averages.

In the first test, we imputed each individual from the SAS and AFR populations with a target-matched imputation panel, that is, SAS (AFR) individuals were imputed using the SAS (AFR) imputation panel. Overall, significantly higher OR per 1SD values were observed on these target-matched imputed sets compared to the EUR imputed set and the non-imputed set (Figure 2A). Next, we looked at the same results when the individuals are split into separate target sets according to their ethnicity. Here too, a higher average OR per 1SD was observed on both SAS and AFR target sets, but only the effect observed for the SAS population reached statistical significance (Figure 2B,C). Most of the results obtained using the other PRS methods had similar trends, although with lower OR per 1SD values (Figure S1).

To reaffirm these results, we used publicly available EUR-predominant GWASs that were not constructed using UKB data to generate PRS models. We chose five traits that had a sufficient number of cases in the UKB SAS and AFR cohorts (see Methods). Using the same pipeline as above, we generated PRS models from the public GWASs and calculated their performance on the SAS and AFR target sets from UKB, imputed with different ethnic panels. The results are summarized in Table 4. For SAS, in two of the three PRS methods, the results obtained using the target-matched imputation panel were slightly better than those obtained using the EUR imputation panel. Surprisingly, for AFR target sets, the PRSs on the non-imputed genotypes outperformed those of the imputed target sets. However, the differences were not statistically significant due to small sample sizes. These results support our previous findings that higher PRS performance may be achieved with a target-matched imputation panel. However, as in our previous analysis, the results for AFR population are more complex.

**Table 4.**
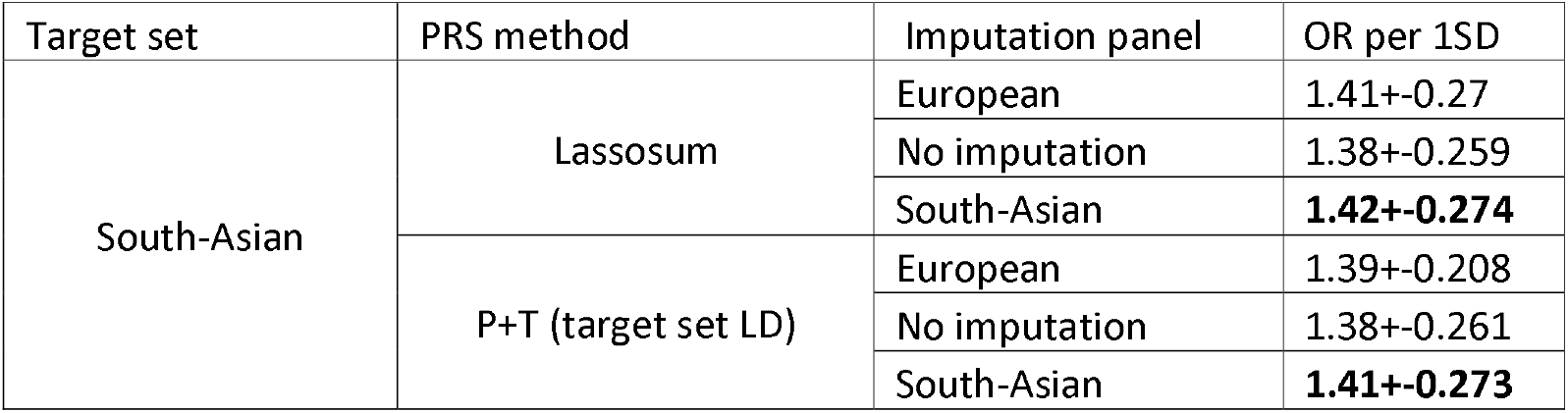
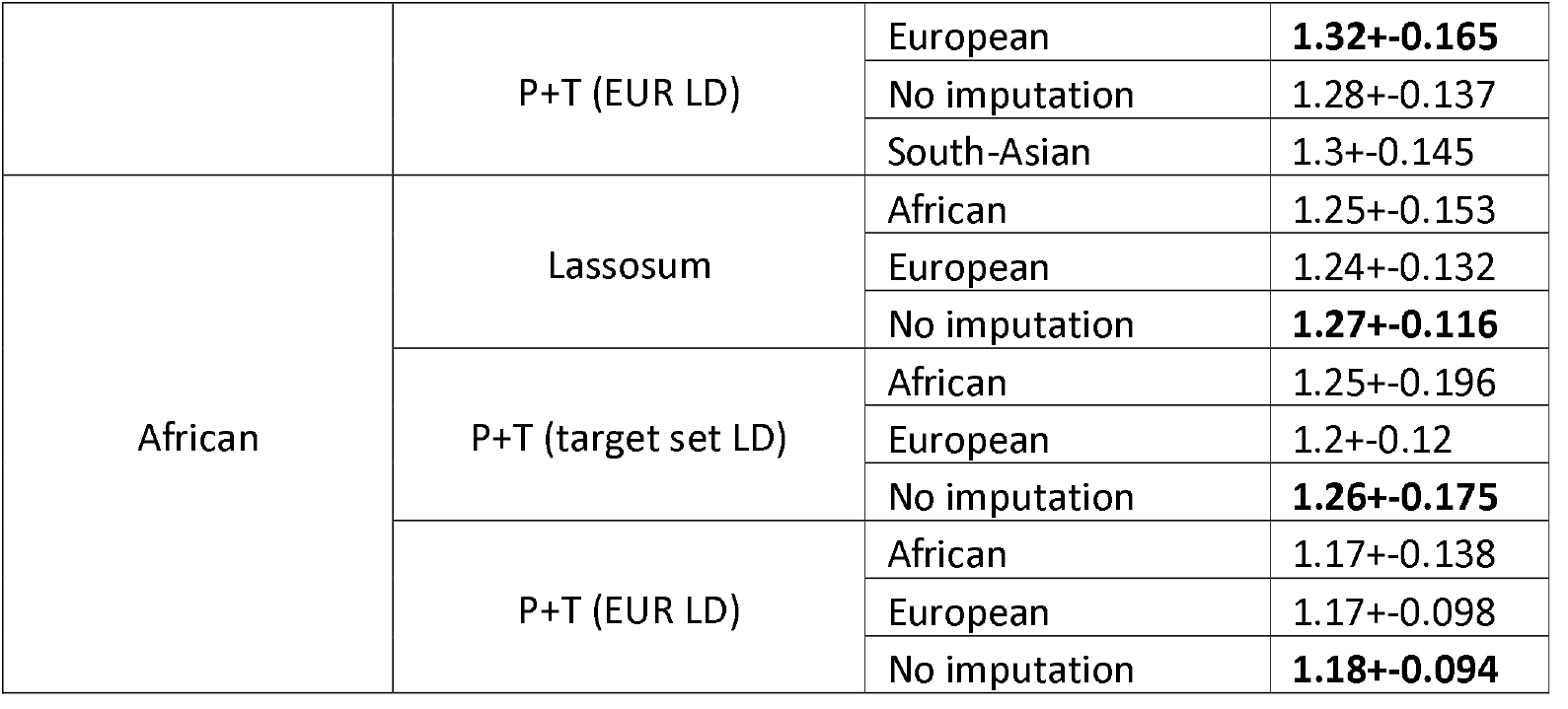
Quality of PRS models on the UKB SAS and AFR cohorts, computed using different imputation panels and PRS methods. PRS were built using EUR-predominant GWASs (n=5) that were not computed with UKB data. P+T: Pruning and thresholding. For P+T, we tested the performance using both EUR LD and the target set LD.

Next, we examined the performance of PRS for Schizophrenia (SCZ), built from EUR SCZ GWAS, on two non-UKB target sets: Ashkenazi Jews (dbGaP access id: phs000448.v1.p1; (Lencz *et al*., 2013); 1044 cases, 2052 controls) and Africans (GAIN; dbGaP access id: phs000021.v3; (Shi *et al*., 2009); 921 cases, 954 controls). We imputed these target sets using three imputation panels from different ethnic groups: EUR, EAS, and AFR. Then, we built EUR GWASs for Schizophrenia (See Methods). Here too, we ran the nested cross-validation scheme to calculate OR per 1SD for each PRS model (See Methods). The results are shown in Table 5. In five out of six cases, the highest performance was obtained when target-matched imputation panels were used.

**Table 5.** OR per 1SD of European Schizophrenia PRS methods for three non-UKB target sets.

| Imputation panel | Method | AFR target set | AJ SCZ target set* |
| --- | --- | --- | --- |
| AFR | P+T (EUR LD) | 1.23±0.06 | 2.13±0.07 |
| EAS |  | 1.44±0.1 | 2.14±0.11 |
| EUR |  | **1.47±0.1** | **2.14±0.1** |
| AFR | P+T (Target LD) | **1.57±0.12** | 2.20±0.11 |
| EAS |  | 1.53±0.13 | 2.10±0.08 |
| EUR |  | 1.33±0.09 | **2.27±0.11** |
| AFR | Lassosum | **1.58±0.12** | 2.58±0.09 |
| EAS |  | 1.57±0.15 | 2.58±0.1 |
| EUR |  | 1.43±0.1 | **2.60±0.08** |

\*Note that for AJ, the EUR imputation panel is the one considered 'target-matched'.

Last, we explored the effect of using different subpopulations of the EUR population as imputation panels. We focused on EUR-based SCZ PRS and imputed the SCZ AJ target set from (Lencz *et al*., 2013) using five imputation panels of EUR sub-populations compiled from the 1KG project (Italy, Spain, Finland, UK, and the US), as well as an imputation panel generated from 100 AJ individuals. OR per 1 SD were calculated by the nested cross-validation scheme as before. In addition to the three PRS methods mentioned above, we also used LDPred2 here. Across all PRS methods, the AJ panel consistently ranked among the top-performing imputation panels, albeit with only marginal improvements (Figure 3).

**Figure 3.**
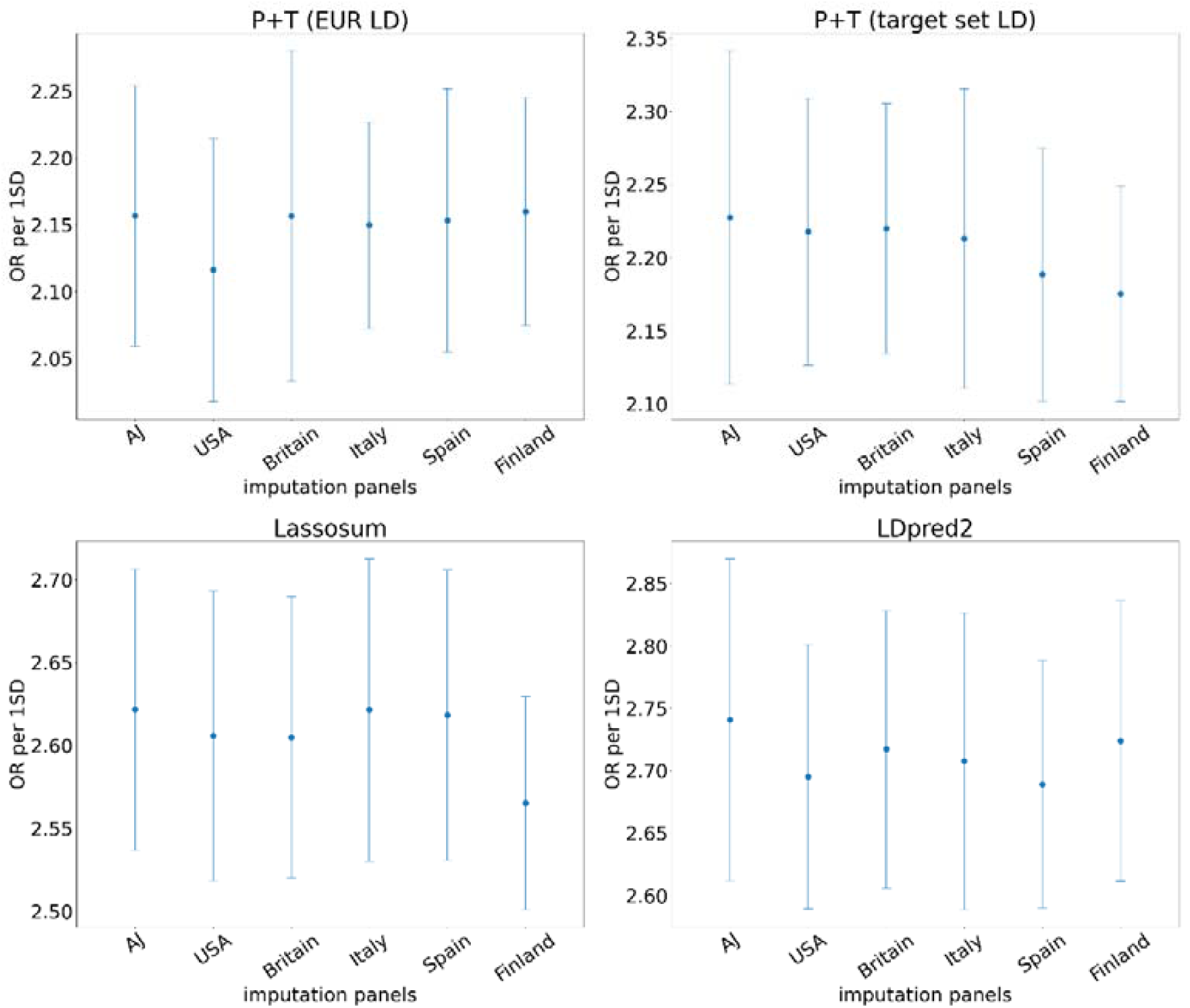
Performance of the EUR-SCZ PRS on the AJ target set imputed using imputation panels derived from different EUR subpopulations. OR per 1SD obtained by several PRS methods using different imputation panels. Dots represent the average value obtained using nested CV, and error bars indicate ±1 sem.

## Discussion

In the emerging era of precision medicine, health disparity has become a major concern. As most available genotypes are predominantly from European population, clinically relevant PRS models are available mainly for that population. Often, many SNPs pertinent to the PRS are missing from the target genotypes. Hence, the imputation process, in which untyped SNPs are completed, could potentially affect the PRS prediction performance.

In this study, we first reconfirmed that imputing genotypes using an ethnic-matched panel is more accurate. However, more accurate SNP imputation does not necessarily imply better risk prediction with PRS. PRS performance deteriorates as the genetic distance between the discovery and the target sets increases (Martin *et al*., 2019). Therefore, when the ethnicity of the target does not match that of the discovery set, the optimal imputation strategy for enhancing PRS performance is not necessarily the one that is optimal for accurately completing missing SNPs.

When utilizing European GWASs derived from UKB data, we observed improved PRS performance primarily when the target set was imputed with an imputation panel from the same ethnic background. This general trend reached statistical significance when tested using Lassosum on AFR and SAS target sets combined, as well as on the SAS target set separately (Figure 2). This trend was consistently observed in two additional independent datasets as well, where higher prediction performance for EUR-PRS was achieved with target-matched imputation panels (Table 5).

While matching the imputation panel to the target population tended to boost PRS performance, the improvement was limited. The statistically significant increase in the OR per 1SD in Lassosum was less than 0.1 on average, suggesting that even when a target-matched imputation panel significantly improves the PRS performance, the magnitude of this effect is moderate. Moreover, when P+T PRS methods were tested, the effect was smaller (Figure S1). Notably, the P+T methods were consistently inferior to Lassosum, which may imply that the benefit of using a target-matched imputation panel is greater when using more advanced PRS methods.

The improvement obtained by using target-matched imputation panel was higher for the SAS population than the AFR one. We suspect that the substantial genetic distance between the EUR population, from which the GWAS was constructed, and the AFR population is the reason why accurately completing missing SNPs did not lead to a significant improvement in EUR-based PRS performance on AFR population.

We analyzed only traits with at least 200 cases in either the AFR or SAS UKB cohorts. Since UKB participants were collected from the general UK population, the numbers of individuals from AFR and SAS ancestries were relatively small (approximately 8,000 and 10,000, respectively). As a result, only twelve disease phenotypes had a sufficient number of cases. As a result, the significance of our analysis was constrained by this limitation. Future studies can focus on continuous traits and thus alleviate this problem.

As most currently available GWASs were conducted on the EUR population, reliable PRSs were developed mostly for EUR individuals. A major effort is currently made to conduct GWASs for non-EUR populations in order to produce clinically relevant PRSs for those ethnicities. This requires collecting large cohorts of cases and healthy individuals for each phenotype and each non-EUR population, which is expensive, logistically complex, and sometimes not feasible for small minorities. Our findings point out that when the target population is moderately close to the EUR population, using an ethnically matched imputation panel has the potential to enhance the prediction performance of European-based PRSs.

## Methods

### Building imputation panels for UKB and GAIN genotypes

We generated imputation panels for the EUR, SAS, EAS, and AFR super-population. White individuals from the US were considered European. For each super-population, we generated a subset of the 1kG sequencing data containing only individuals from that population. Then, for each subset, we used SHAPEIT2 (Delaneau *et al*., 2011) to generate its corresponding imputation panel.

### Building imputation panels for analyzing AJ imputation panel performance

To build an imputation panel for AJ, we used WGS data of 100 AJ individuals from (Carmi *et al*., 2014). In analyses that compared the performance of AJ imputation panel to other panels (Table 2, Figure 3), we generated imputation panels comprising only SNPs that were found in both the AJ dataset and 1kG. To do so, we used BCFTOOLS (Danecek *et al*., 2021) to annotate the SNP in the AJ dataset with SNP ids (rsid) from the 1kg dataset. In this process, 8,637,756 SNPs in the AJ dataset were assigned with rsid. We kept only those SNPS (i.e., that were shared between the AJ and 1kG datasets). Then, we used SHAPEIT2 (Delaneau *et al*., 2011) to phase the remaining SNPs in the AJ dataset. Finally, we used SHAPEIT2 to generate imputation panels.

### Genotype imputation

Using the imputation panels we generated (see above), we imputed genotypes from the UKB, GAIN (O’Donovan *et al*., 2008), and the SCZ-AJ datasets (Lencz *et al*., 2013). Imputation was done using IMPUTE2 (Howie *et al*., 2011).

### Nested CV scheme

We applied a variant of the nested CV scheme described in (Levi et al., 2023). Briefly, we split each target set cohort into six sets. Next, we held out one set and used the other five sets to perform a standard 5-fold CV, in which four out of five parts are used to derive PRS models with different predefined sets of hyper-parameters, and then the resulting models are applied on the 5th part. After iterating over the five combinations of training and test sets, we chose the best performing hyper-parameter set. Finally, we applied the resulting PRS model on the held-out set. We repeated this entire process six times, each with a different hold-out set and took the average.

### PRS methods

We used four PRS methods: (1) P+T (EUR LD): Using PLINK, we clumped the GWAS results according to LD in the EUR population, and then we filtered the remaining SNPs based on a significance threshold. (2) P+T (Target LD): Here, when applying LD clumping in PLINK, we used LD inferred from the training set. The rest is similar to the previous method. (3) Lassosum: we generated a PRS model using a reference panel calculated from genotype data (i.e., the training set). (4) LDPred2 (grid mode). We supplied LDpred2 with a training set that comes from the same population as the target set. As LDPred2 is computationally intensive, we used it only in our last analysis on EUR-SCZ PRS (Figure 3).

### Generating GWAS summary statistics

For GWASs generated from the UKB, we considered only phenotypes that had at least 200 cases in either the AFR or the SAS target sets in UKB (Table S3). For each phenotype, we generated GWAS summary statistics by applying plink’s --assoc command to the imputed genotypes of the EUR population provided by the UKB.

### GWAS summary statistics from public datasets

For the analysis of non-UKB EUR GWAS (Table 4), we used publicly available EUR-predominant GWAS for five traits that have at least 200 AFR and SAS cases in UKB: Systolic Blood Pressure (BP) (Evangelou *et al*., 2018), Cholesterol levels (Willer *et al*., 2013), Type 2 Diabetes (Mahajan *et al*., 2018), Gastric Reflux (An *et al*., 2019), and Major Depression (Howard *et al*., 2019). For the analysis of SCZ PRS on non-UKB target sets (Table 5, Figure 3), we used a leave-one-out version of the SCZ GWAS from (Ripke *et al*., 2014), where individuals presented in the AJ target set were excluded.

## Data Availability

All data produced in the present study are available upon reasonable request by the UK Biobank and dbGAP and the Human Genome Phenome Archive.

## Acknowledgments

We thank Todd Lencz for providing us with the leave-one-out version of the SCZ GWAS and for his insightful comments.

This research has been conducted using the UK Biobank application no. 56885.

R.E. is a Faculty Fellow of the Edmond J. Safra Center for Bioinformatics, Tel Aviv University. This work was carried out in partial fulfillment of the requirements for the Ph.D. degree of H.L. at the Blavatnik School of Computer Science, Tel Aviv University.

## Supplementary Information

**Table S1.** Ethnic groups from the 1kG that comprise the super populations in the analyses described in Tables 1-3,S2-3.

| EUR | SAS | EAS | AFR | AFR2 |
| --- | --- | --- | --- | --- |
| GBR (British from England and Scotland) | BEB (Bengali in Bangladesh) | CDX (Chinese Dai in Xishuangbanna, China) | ACB (African Caribbean in Barbados) | MSL (Mende in Sierra Leone) |
| FIN (Finnish in Finland) | GIH (Gujarati Indians in Houston, Texas, USA) | CHB (Han Chinese in Beijing, China) | ESN (Esan in Nigeria) | ASW (African Ancestry in SW USA) |
| TSI (Toscani in Italia) | ITU (Indian Telugu in the U.K.) | CHS (Han Chinese South) | GWD (Gambian in Western Division – Mandinka) |  |
| IBS (Iberian populations in Spain) | PJL (Punjabi in Lahore, Pakistan) | JPT (Japanese in Tokyo, Japan) | LWK (Luhya in Webuye, Kenya) |  |
| CEU (Utah residents (CEPH) with northern and western European ancestry) | STU (Sri Lankan Tamil in the UK) | KHV (Kinh in Ho Chi Minh City, Vietnam) | YRI (Yoruba in Ibadan, Nigeria) |  |

**Table S2. See the file Table S2.xlsx**

**Table S3. See the file Table S3.xlsx**

**Table S4.** 12 Traits used to construct GWASs from the UKB genotypes. All traits are binary. Cases were defined according to a diagnosis given to the individual’s doctor (self-reported).

| Trait | Number of AFR cases | Number of SAS cases |
| --- | --- | --- |
| Angina | 206 | 531 |
| Asthma | 969 | 1178 |
| High cholesterol | 894 | 1868 |
| Cataract | 236 | 351 |
| gastro-esophageal reflux (GERD) / gastric reflux | 249 | 436 |
| Hay fever | 493 | 546 |
| Hypertension | 2984 | 2864 |
| Hypothyroidism/Myxedema | 172 | 584 |
| Major depression disorder | 254 | 389 |
| Osteoarthritis | 405 | 517 |
| Type 2 Diabetes | 792 | 1520 |
| Urinary tract infection | 479 | 119 |

**Table S5.** OR per 1SD for three non-UKB target sets. PRSs were generated from EAS GWASs. Lassosum was tested with two values for the parameter LDBlocks: EUR and ASN (hg19)

| Imputation panel | Method | AFR target set |  | AJ SCZ target set |  |
| --- | --- | --- | --- | --- | --- |
|  |  | OR per 1SD | SEM | OR per 1SD | SEM |
| AFR | P+T (EUR LD) | **1.11** | 0.02 | **0.97** | 0.04 |
| EAS |  | 0.94 | 0.05 | 0.94 | 0.02 |
| EUR |  | 1.00 | 0.02 | 0.96 | 0.03 |
| EAS | P+T (Target LD) | 0.99 | 0.04 | 1.02 | 0.04 |
| SAS |  | 1.07 | 0.04 | 1.01 | 0.03 |
| EUR |  | **1.09** | 0.06 | **1.04** | 0.04 |
| AFR | Lassosum (EUR) | 0.98 | 0.03 | 1.07 | 0.05 |
| EAS |  | 1.00 | 0.03 | **1.10** | 0.05 |
| EUR |  | **1.08** | 0.07 | 1.08 | 0.04 |
| AFR | Lassosum (ASN) | 1.00 | 0.02 | 1.04 | 0.05 |
| EAS |  | 0.99 | 0.02 | **1.11** | 0.05 |
| EUR |  | **1.08** | 0.07 | 1.07 | 0.04 |

**Figure S1.**
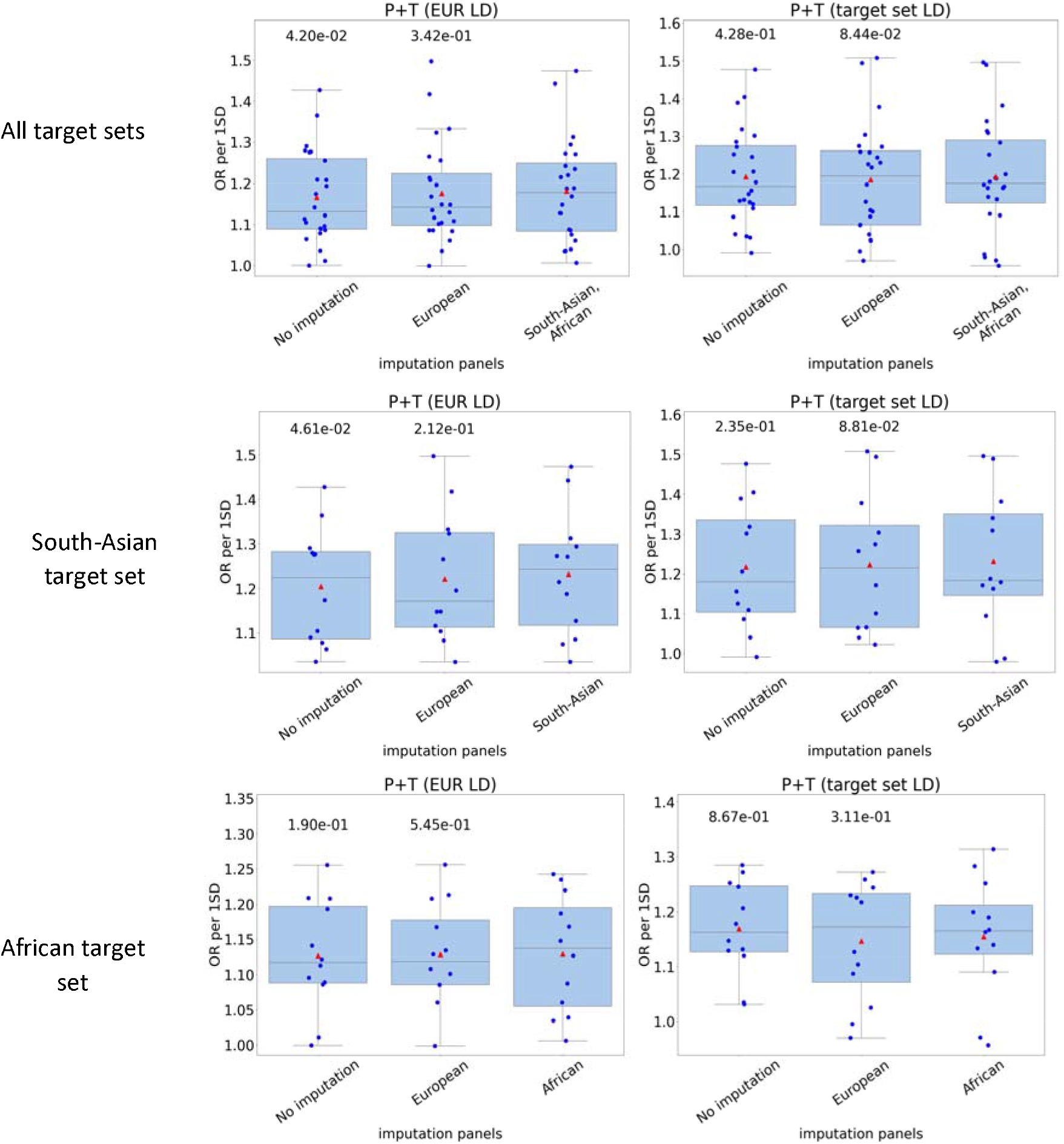
The effect of ethnic composition of the imputation panels on the performance of P+T (EUR LD) and P+T (target set LD) PRS methods when applied to target set of different ethnicity than the GWAS. The graphs show OR per 1SD of 12 traits. PRSs were built from GWASs computed on UKB EUR individuals. Results are shown for (A) both SAS and AFR (B) SAS only (C) AFR only. The p-values above each boxplot compare the results with the imputation panel listed below it to the results with the imputation panel of the target population. P-values were calculated using one-tailed Wilcoxon test. Red triangles are the averages.

